# Impact of COVID-19 pandemic on the socioeconomic inequality of health behavior among Japanese adolescents: a two-year-repeated cross-sectional survey

**DOI:** 10.1101/2022.08.11.22278499

**Authors:** Akira Kyan, Minoru Takakura

## Abstract

**Background:** Although disparities in socioeconomic status in health behaviors have been highlighted globally, they are not well understood in Japanese adolescents. The purpose of this study was to clarify the changes in socioeconomic disparities in adolescents’ fundamental health behaviors, such as physical activity, screen time (ST), sleep, breakfast intake, and bowel movement before and during COVID-19.

**Methods:** This was a repeated cross-sectional study which used data from the 2019 and 2021 National Sports-Life Survey of Children and Young in Japan. Data of 766 and 725 participants in 2019 and 2021, respectively, were analyzed. Favorable health behaviors were defined as daily moderate-to-vigorous physical activity (MVPA) of at least 60 minutes, ST of less than 2 hours, sleep of 8 to 10 hours, daily breakfast intake, and bowel movement frequency of at least once in every 3 days. We calculated the slope index of inequality (SII) and relative index of inequality (RII) in each health behavior for equivalent household income levels for assessing absolute and relative economic inequalities.

**Results:** Compliance with MVPA and ST recommendation significantly declined from 20.1% and 23.0% in 2019 to 11.7% and 14.9% in 2021, respectively. The SII and RII increased in MVPA for income levels, but decreased in daily breakfast in 2019 to 2021. Although the widening and narrowing of the disparity was inconclusive for ST, it exacerbated for the higher income groups.

**Conclusions:** Our study revealed widening of economic disparities in the achievement of recommended MVPA and narrowing of it in breakfast intake among adolescents before and during COVID-19.

## Introduction

Socioeconomic disparities of health in Japan have become an issue within the last 20 years after rapid economic growth. A systematic review of health disparities reported that the gradient of disparities was modest in the first decades since the 1990s compared to the Western countries.^1^ As the situation worsened, it was recognized as a public health concern.^2^ Since 2013, Japan’s health policies have tried to correct this situation.^3^

In Western countries, disparities by a family or neighborhood’ economic status have been observed in several fundamental health behaviors, such as physical activity (PA),^4^ sedentary behavior (screen time (ST)),^5^ sleep behavior,^6^ bowel movement frequency (constipation),^7^ and food intake,^8^ among adolescents. In contrast, they are not well understood in Japan as it has only been a few decades since health inequalities first gained attention. In recent examples of inequality of health behavior, daily breakfast intake was higher in the high-income group,^9^ while socioeconomic disparities in food and nutrient intake were restrained due to school lunches on weekdays.^10^ There are no studies on the relationship between socioeconomic status (SES) and exercise in Japanese adolescents, except for one inconclusive study in a local journal.^11^

During the COVID-19 pandemic, fundamental health behaviors deteriorated among adolescents worldwide.^12–14^ National surveys among Japanese adolescents reported that exercise, which included active play, declined significantly, screen time of ≥ 2 hours per day increased significantly, and daily breakfast intake, adequate sleep of 8-10 hours, and bowel movement frequency remained almost unchanged.^15,16^ The COVID-19 pandemic disrupted both people’s healthcare and lifestyle, social, and economic activities. In Japan, to prevent the transmission of the virus, policies restricted people’s activities, such as restaurants closures and restrictions on large-scale events.^17^ Thereby, the service industry experienced a significant drop in sales, and many businesses were forced into unemployment or bankruptcy.^17^ In contrast, the information and communication industry, which included online businesses, saw increased sales.^17^ Hence, the accompanying widening of household income inequality became a concern. A public health concern was that this may increase disparities in people’s health or health behaviors.^18^ The possibility of increased disparities in mental well-being ^19^ and recreational PA ^20^ among Japanese adults was also recently reported. Regarding dietary intake among Japanese adolescents, a recent retrospective study found that meat, fish, eggs, and vegetables intake worsened, especially in low-income households, during school closure (from April 16 to May 6, 2020). Although, it mended after school lunch restarted.^21^ However, no study has been conducted on other health behaviors in Japan or globally.

Adolescent health behaviors have been documented to determine physical and mental growth and development and health during adolescence, and also how these are carried into the future.^22^ It is a right for children to have a healthy life.^23^ Those suffering from inequality due to their a home environment beyond their control are victims. While some health behaviors, affected by the COVID-19 pandemic, should be recovered for adolescents, understanding how these behaviors vary with household income is important to create intervention strategies (to provide the necessary support) and policies. Thus, this study aimed to clarify the trends in socioeconomic disparities in adolescents’ health behaviors, especially fundamental health behaviors, such as PA, ST, breakfast intake frequency, and bowel movement frequency before and during the COVID-19 pandemic.

## Material and Methods

### Data source

This study used data from the 2019 and 2021 National Sports-Life Survey of Children and Young People conducted by the Sasakawa Sports Foundation (SSF).^16^ Nationwide cross-sectional surveys have been conducted every two years since 2001. From the 2017 survey, the target age groups were categorized as 4-11 and 12-21 years. The survey mainly measured exercise and sports participation of children and youth during after-school periods and holidays, sports environments, and health behaviors, which included sleep time, media usage time, and bowel movement frequency. Data were collected via self-administered questionnaires from adolescents and parents/guardians between June and July of each survey year.

Each year’s study sample were selected using a two-staged stratified random sampling method from 225 locations (2019: 61 metro areas, 94 and 50 cities with a population of more than 100,000 and less than 100,000, respectively, and 20 towns/villages; 2021: 61 metro areas, 96 and 50 cities with a population of more than 100,000 and less than 100,000, respectively, and 18 towns/villages). These were proportionally distributed from the strata by district/city size based on the population of the Basic Resident Register as of January 1^st^ in the year prior to each survey. The study area established during the 2015 census was used as the primary sampling unit at the time of the survey. The target sample size allocated to each location was 10-19. Survey points were extracted by the probability proportional sampling method to calculate the sampling interval for strata in which two or more survey points were assigned using the following formula:

#### Total number of people aged 4-21 years in the study area in the strata/number of survey sites calculated in strata

The survey included 3,000 individuals. Detailed information on the survey methods can be found on the SSF website.^16^

We used data from the surveys of young people aged 12-21 years in 2019 and 2021. The number of responses for 2019 and 2021 were 1,675 (response rate 55.8%) and 1,663 (55.4%), respectively. The analysis included 12-to 18-year-olds and excluded 18-year-olds who did not attend high school. In the Japanese educational system, most students enrolled in high school were 15-to 18-year-olds. Of these, most 17- and 18-year-olds were enrolled in the third grade and shared school routines. This was the reason we included 18-year-olds who attended high school, although the age for the analysis of PA, ST, and sleep recommendations was 17 years. The number of participants who met the inclusion criteria of age and school enrollment was 1076 (570 boys and 506 girls) and 1025 (517 boys and 508 girls) for 2019 and 2021, respectively. After individuals with missing variables were excluded, data from 766 participants (359 boys and 407 girls) in 2019 and 725 participants (365 boys and 360 girls) in 2021 were analyzed. Ethical approval was not required as this study was a secondary analysis conducted using public datasets from the SSF that did not include identifiable personal information.

### Measures

#### Physical activity

PA was measured using the Japanese version of the Health Behavior in School-aged Children survey (HBSC) questionnaire.^24^ The HBSC questionnaire was widely used as the basis of moderate-to-vigorous PA (MVPA) surveillance in adolescents.^25^ PA referred to any activity that increased heart rate and made an individual feel out of breath for some time. It included sports, school activities, playing with friends, or walking to school. Examples included running, brisk walking, rollerblading, biking, dancing, skateboarding, swimming, playing soccer, basketball, football, surfing, etc. The question item was “over the past seven days, on how many days were you physically active for a total of at least 60 minutes per day?.” Participants were dichotomized into either active or inactive based on whether they achieved seven days per week of 60 minutes MVPA, according to the World Health Organization (WHO) guidelines on PA and sedentary behavior.^26^

#### Screen time

ST was assessed by enquiring regarding recreational TV/DVD viewing time and computer/game/smartphone usage on weekdays and weekends separately. The question was “for how many hours do you watch TV or DVD or use computer, video games (including TV, computer, and cellular device games, etc.) or use a smartphone per day outside of school and/or work?.” Possible answers were “less than half-hour/day,” “half-hour to 1 h/day,” “1–2 h/day,” “2–3 h/day,” “3–4 h/day,” “4–5 h/day,” “more than 5 h/day,” and “I do not know.” The answers were recorded in minutes using the midpoint method. “I do not know” was deemed as a missing case. Average minutes of daily ST were weighted by weekdays and weekends using the following formula:

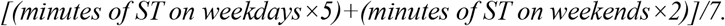

We categorized the participants based on the cut-off of 2 h of recreational ST ^26^. Although there are no standardized ST questionnaire items, many previous studies have adopted items corresponding to the time spent watching TV and using smartphones, tablets, and PCs as indicators, which is comparable to our study.^27^

#### Sleep duration

Sleep duration was assessed by questioning bedtime and awakening hours on weekdays and weekends separately. Participants were asked to report the times they typically slept at night and woke up in the morning on weekdays and weekends. Sleep duration was calculated by subtracting wake-up time from bedtime.

Average minutes of daily sleep were weighted by weekdays and weekends using the following formula:

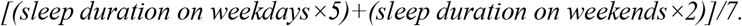

Participants were grouped based on whether their sleep duration was within the range (8.0–10.0 h/night) recommended by the National Sleep Foundation ^31^.

#### Breakfast

Breakfast frequency was determined via the question “how often do you have breakfast per week?” The possible responses were “almost every day,” “ate 4-5 days,” “ate 2-3 days,” and “very few.” Participants were dichotomized into almost every day or others based on the recommendation of the Ministry of Agriculture, Forestry and Fisheries, Japan ^32^.

#### Bowel movement frequency

Bowel movement frequency was determined via the question “how often do you have bowel movement?” The possible responses were “almost every day,” “once per two days,” “once per three days,” “less than once per three days,” and “irregularly.” We dichotomized participants into two groups: “almost every day to once every three days” and “less than once every three days and irregular” as per the definition of constipation ^7^.

The aforementioned measurements for PA, ST, sleep duration, breakfast, and bowel movement frequency are similar to those used in international epidemiology surveys ^7,25,28^ and national surveys in Japan.^33^

#### Household Income

Participants’ parents/guardians were asked regarding their annual pre-tax household income using 11 options that ranged from “no income” to “10 million yen or more.” “I do not know” was deemed as a missing case. The midpoint of each option was substituted. The equivalent household income was calculated by dividing household income by the square root of the number of household members ^34^. It was categorized based on one-half of the median equivalent household income as a cut-off point: “less than 1.375 million yen,” “1.375 to less than 2.75 million yen,” and “2.75 million yen or more” ^35^. One million yen was roughly 12,000 US dollars at the time of the survey.

#### Covariate

Place of residence, family structure, sex, age, sports participation, self-rated health, and preference of PA were used as covariates and considered as potential confounders ^4,36^. Place of residence was categorized into two groups based on the population size, with a cut-off point of 100,000 individuals. Family structure was assessed using information regarding the people living together, coded as “lived with both parents” or “other.” Sports participation was assessed by asking whether the participants were involved in extra-curricular exercise activity in school, sports clubs in local community, or privately. Self-rated health was dichotomized into “good” and “poor.” Preference for PA was dichotomized into “liked” and “disliked.”

### Statistical analysis

The percentage of compliance with health behavior recommendation was estimated by each income level and each survey year. Cochran-Armitage test for trend was used to assess time trends in the percentage of compliance with each recommendation of health behavior and prevalence of constipation by each income level. Fisher’s exact test was performed to evaluate differences in compliance and prevalence of constipation by each income level and survey year. Socioeconomic inequalities in health behavior between low- and high-income groups were assessed by absolute and relative measures, estimated with 95% confidence intervals (CIs) for income levels in each survey year. For the absolute measures, the slope index of inequality (SII) ^37–39^ was calculated using generalized linear models with binomial distribution and identity link function. The coefficient yielded an estimate of the absolute inequality. When this binomial model failed to converge, a generalized linear model with normal distribution and identity link function was used.^40^ For the relative measures, the relative index of inequality (RII) ^37–39^ was calculated using generalized linear models with binomial distribution and log link function. The exponentiated coefficient yielded an estimate of the relative inequality. The SII and RII were estimated using ridit score for income levels as an independent variable. They were summary measures of inequality as the changes in health behavior between the bottom and top points in the income hierarchy while considering the cumulative distribution in each income level.^41^ The SII estimated the absolute predicted difference in health behavior between the theoretical highest and lowest income level, interpreted as the difference in appearance probability of the health behavior at the two extremes of the socioeconomic spectrum, while RII refers to the ratio of the appearance probabilities. Time trends of the measures were assessed by the inclusion of the interaction term between income levels (ridit scores) and survey year.^41^ In the model, survey year was treated as a continuous variable coded as 1 for 2019 and 2 for 2021.^41^ Wald test was used to evaluate if the interaction term was statistically significant. These models were adjusted the confounding factors.

## Results

Table 1 shows the distribution of the participants by sociodemographic characteristics and survey years. There were significant relationships between survey year and MVPA and ST (*p*<0.05). The number and percentage of missing each variable are shown in the e-Table (appendix). The missing percentages for each variable were similar in both surveys. Household income was missing by approximately 22% in each survey as “I do not know” was treated as a missing value. Among the subjects who met the age inclusion criteria, there were no significant differences in demographic factors such as age, sex, or residence area between those who were excluded from the analysis due to missing data and those who were included in the analysis (data not shown). For the interest variables, differences in income and PA only presented in 2019. Individuals with a lower income and those engaging in non-compliance with PA recommendations were more likely to be excluded.

**Table 1.** Distribution of the study participants by sociodemographic characteristics

|  | 2019 |  | 2021 |  | P<sup>a</sup> |
| --- | --- | --- | --- | --- | --- |
|  | N | % | N | % |  |
| Total | 766 |  | 725 |  |  |
| Sex |  |  |  |  |  |
| Male | 359 | 46.9 | 365 | 50.3 | 0.195 |
| Female | 407 | 53.1 | 360 | 49.7 |  |
| Age group (year) |  |  |  |  |  |
| 12 | 88 | 11.5 | 78 | 10.8 | 0.571 |
| 13 | 152 | 19.8 | 129 | 17.8 |  |
| 14 | 122 | 15.9 | 110 | 15.2 |  |
| 15 | 140 | 18.3 | 120 | 16.6 |  |
| 16 | 119 | 15.5 | 133 | 18.3 |  |
| 17 | 115 | 15.0 | 120 | 16.6 |  |
| 18 | 30 | 3.9 | 35 | 4.8 |  |
| Residence area |  |  |  |  |  |
| 21 metropolises and cities with population of 100,000+ | 679 | 88.6 | 652 | 89.9 | 0.452 |
| Cities with population < 100,000, and towns and villages | 87 | 11.4 | 73 | 10.1 |  |
| Equivalent household income (million yen) |  |  |  |  |  |
| < 1.375 | 69 | 9.0 | 66 | 9.1 | 0.347 |
| 1.375 - 2.75 | 259 | 33.8 | 220 | 30.3 |  |
| 2.75+ | 438 | 57.2 | 439 | 60.6 |  |
| Family structure |  |  |  |  |  |
| Both parents | 651 | 85.0 | 635 | 87.6 | 0.153 |
| Other | 115 | 15.0 | 90 | 12.4 |  |
| Sports participation |  |  |  |  |  |
| No | 270 | 35.2 | 292 | 40.3 | 0.048 |
| Yes | 496 | 64.8 | 433 | 59.7 |  |
| Self-rated health |  |  |  |  |  |
| Good | 650 | 84.9 | 602 | 83.0 | 0.359 |
| Bad | 116 | 15.1 | 123 | 17.0 |  |
| Preference of physical activity |  |  |  |  |  |
| Dislike | 142 | 18.5 | 130 | 17.9 | 0.789 |
| Like | 624 | 81.5 | 595 | 82.1 |  |
| MVPA |  |  |  |  |  |
| < 7 days | 612 | 79.9 | 640 | 88.3 | 0.000 |
| 7 days | 154 | 20.1 | 85 | 11.7 |  |
| Screen time |  |  |  |  |  |
| < 2 h | 176 | 23.0 | 108 | 14.9 | 0.000 |
| ≥ 2 h | 590 | 77.0 | 617 | 85.1 |  |
| Adequate sleep duration |  |  |  |  |  |
| 8hr–10hr | 484 | 63.2 | 469 | 64.7 | 0.553 |
| Other | 282 | 36.8 | 256 | 35.3 |  |
| Breakfast |  |  |  |  |  |
| Almost everyday | 126 | 16.4 | 121 | 16.7 | 0.944 |
| Other | 640 | 83.6 | 604 | 83.3 |  |
| Constipation |  |  |  |  |  |
| Yes | 77 | 10.1 | 76 | 10.5 | 0.798 |
| No | 689 | 89.9 | 649 | 89.5 |  |

Note: <sup>a</sup> $\chi^2$ test

Table 2 shows the prevalence in compliance with each health behavior recommendation by income levels and survey years. Overall, compliance with PA and ST recommendation significantly declined from 20.1% and 23.0% in 2019 to 11.7% and 14.9% in 2021, respectively. Compliance with sleep and breakfast frequency recommendation and constipation prevalence remained unchanged between 2019 (63.2%, 83.6%, and 10.1%, respectively) and 2021 (64.7%, 83.3%, and 10.5%, respectively). While declining trends in PA were observed for all income levels, declining trends in ST were observed for only the high-income level. Regarding breakfast consumption, no overall change was observed between 2019 and 2021. However, a non-statistically significant declining trend was observed only at the high-income level. Table 2 also shows the results of Fisher’s exact test. A clear difference between 2019 and 2021 emerged in breakfast intake: the percentage was lower in the low-income group and higher in the highest-income group in 2019 (*p*<0.001). However, the differences were obscured in 2021 (*p*=0.151). A similar trend was observed for ST, although it was not statistically significant (*p*=0.097 and 0.862 in 2019 and 2021, respectively). PA showed differences by income levels in 2021, although it was not statistically significant (*p*=0.063); however, not in 2019 (*p*=0.907). These differences by socioeconomic levels were not found in compliance with sleep duration (p=0.916 and 0.356 in 2019 and 2021, respectively) and bowel frequency (*p*=0.503 and 0.265 in 2019 and 2021, respectively).

**Table 2.** Prevalence of the health behavior in 2019 and 2021 according to the equivalent household income

|  | Compliance with MVPA guideline |  |  |  |  | Compliance with screen time guideline |  |  |  |  | Compliance with sleep duration guideline |  |  |  |  | Daily breakfast intake |  |  |  |  | Constipation <sup>a</sup> |  |  |  |  |
| --- | --- | --- | --- | --- | --- | --- | --- | --- | --- | --- | --- | --- | --- | --- | --- | --- | --- | --- | --- | --- | --- | --- | --- | --- | --- |
|  | 2019 |  | 2021 |  |  | 2019 |  | 2021 |  |  | 2019 |  | 2021 |  |  | 2019 |  | 2021 |  |  | 2019 |  | 2021 |  |  |
|  | n | (%) | n | (%) | P<sup>b</sup> for trend | n | (%) | n | (%) | P<sup>b</sup> for trend | n | (%) | n | (%) | P<sup>b</sup> for trend | n | (%) | n | (%) | P<sup>b</sup> for trend | n | (%) | n | (%) | P<sup>b</sup> for trend |
| Total | 154 | 20.1 | 85 | 11.7 | <0.001 | 176 | 23 | 108 | 14.9 | <0.001 | 282 | 36.8 | 256 | 35.3 | 0.545 | 640 | 83.6 | 604 | 83.3 | 0.901 | 77 | 10.1 | 76 | 10.5 | 0.784 |
| Equivalent household income (million yen) |  |  |  |  |  |  |  |  |  |  |  |  |  |  |  |  |  |  |  |  |  |  |  |  |  |
| < 1.375 | 15 | 21.7 | 3 | 4.5 | 0.003 | 14 | 20.3 | 9 | 13.6 | 0.304 | 27 | 39.1 | 19 | 28.8 | 0.205 | 48 | 69.6 | 50 | 75.8 | 0.420 | 5 | 7.2 | 4 | 6.1 | 0.782 |
| 1.375 - 2.75 | 53 | 20.5 | 22 | 10.0 | 0.002 | 49 | 18.9 | 35 | 15.9 | 0.388 | 95 | 36.7 | 84 | 38.2 | 0.735 | 208 | 80.3 | 189 | 85.9 | 0.105 | 30 | 11.6 | 28 | 12.7 | 0.702 |
| 2.75+ | 86 | 19.6 | 60 | 13.7 | 0.018 | 113 | 25.8 | 64 | 14.6 | <0.001 | 160 | 36.5 | 153 | 34.9 | 0.604 | 384 | 87.7 | 365 | 83.1 | 0.058 | 42 | 9.6 | 44 | 10.0 | 0.829 |
| P for Fisher's exact test | 0.907 |  | 0.063 |  |  | 0.097 |  | 0.863 |  |  | 0.916 |  | 0.356 |  |  | <0.001 |  | 0.151 |  |  | 0.503 |  | 0.265 |  |  |

Note: MVPA moderate-to-vigorous physical activity,
<sup>a</sup> bowel movement frequency is less than once every 3 days.
<sup>b</sup> Cochran-Armitage trend test

Table 3 and e-graph (appendix) show the SII and RII in each health behavior for the income levels by survey years. Significant increase and decrease trends of disparities were seen in compliance of MVPA and daily breakfast intake. The SII in MVPA increased from -2.37% (95%CI 13.62–8.87) in 2019 to 11.2% (95%CI 2.90–19.50) in 2021. Although it was not statistically significant in the crude model (*p*=0.057), the adjusted model showed significant increase trends (*p*=0.032). The RII in MVPA increased from 0.86 (95%CI 0.43–1.73) in 2019 to 3.03 (95%CI 1.16–7.93) in 2021 (*p*=0.038). In contrast, the SII in breakfast intake decreased from 20.22% (95%CI 9.4–31.04) in 2019 to 1.35% (95%CI -9.87–12.57) in 2021 (*p*=0.018). The RII also decreased from 4.34 (95%CI 2.05–9.18) in 2019 to 1.11 (95%CI 0.52.47) in 2021 (*p*=0.015). The SII in ST decreased from 20.72% (95%CI 7.71–33.72) in 2019 to 4.74% (95%CI - 7.97–17.45) in 2021, although it was not statistically significant (*p*=0.085). Similarly, for RII, the disparity decreased numerically, although it was not statistically significant. For sleep duration and bowel movement, no disparities or changing trends were observed. Except the SII in PA, all adjusted models showed almost similar results as the crude models.

**Table 3.** Slope index of inequality (SII) and relative index of inequality (RII) in the health behavior according to the income levels by survey year

|  | 2019 |  | 2021 |  | P for trend <sup>c</sup> |
| --- | --- | --- | --- | --- | --- |
| MVPA |  |  |  |  |  |
| Crude SII (95% CI) | -2.37 | (-13.62, 8.87) | 11.20 | (2.9, 19.5) | 0.057 |
| Adjusted SII<sup>a</sup> (95% CI) <sup>b</sup> | -7.94 | (-18.94, 3.06) | 8.86 | (0.23, 17.48) | 0.032 |
| Crude RII | 0.86 | (0.43, 1.73) | 3.03 | (1.16, 7.93) | 0.038 |
| Adjusted RII<sup>a</sup> (95% CI) | 0.59 | (0.28, 1.23) | 2.50 | (0.95, 6.58) | 0.018 |
| Screen time |  |  |  |  |  |
| Crude SII (95% CI) | 20.72 | (7.71, 33.72) | 4.74 | (-7.97, 17.45) | 0.085 |
| Adjusted SII<sup>a</sup> (95% CI) <sup>b</sup> | 15.85 | (3, 28.7) | 6.42 | (-6.19, 19.03) | 0.137 |
| Crude RII | 2.57 | (1.4, 4.7) | 1.27 | (0.66, 2.47) | 0.125 |
| Adjusted RII<sup>a</sup> (95% CI) | 2.16 | (1.13, 4.12) | 1.41 | (0.71, 2.82) | 0.208 |
| Sleep duration |  |  |  |  |  |
| Crude SII (95% CI) | -1.84 | (-15.32, 11.64) | -0.15 | (-13.96, 13.66) | 0.864 |
| Adjusted SII<sup>a</sup> (95% CI) <sup>b</sup> | -5.42 | (-18.68, 7.83) | 0.40 | (-12.42, 13.23) | 0.707 |
| Crude RII | 0.92 | (0.52, 1.65) | 0.99 | (0.54, 1.82) | 0.865 |
| Adjusted RII<sup>a</sup> (95% CI) | 0.79 | (0.42, 1.46) | 1.02 | (0.52, 1.99) | 0.720 |
| Breakfast |  |  |  |  |  |
| Crude SII (95% CI) | 20.22 | (9.4, 31.04) | 1.35 | (-9.87, 12.57) | 0.018 |
| Adjusted SII<sup>a</sup> (95% CI) <sup>b</sup> | 18.98 | (8.4, 29.56) | 1.29 | (-9.44, 12.01) | 0.018 |
| Crude RII | 4.34 | (2.05, 9.18) | 1.11 | (0.5, 2.47) | 0.015 |
| Adjusted RII<sup>a</sup> (95% CI) | 4.19 | (1.89, 9.27) | 1.14 | (0.5, 2.62) | 0.022 |
| Constipation |  |  |  |  |  |
| Crude SII (95% CI) | -1.09 | (-9.29, 7.11) | -0.82 | (-9.46, 7.82) | 0.965 |
| Adjusted SII<sup>a</sup> (95% CI) <sup>b</sup> | 2.68 | (-5.61, 10.97) | 3.08 | (-5.44, 11.6) | 0.865 |
| Crude RII | 0.89 | (0.36, 2.19) | 0.92 | (0.37, 2.29) | 0.960 |
| Adjusted RII<sup>a</sup> (95% CI) | 1.49 | (0.51, 4.31) | 1.44 | (0.53, 3.89) | 0.845 |

Note: SII slope index of inequality (%), RII relative index of inequality, CI confidence interval
<sup>a</sup> Adjusted for place of residence, family structure, sex, age, sports participation, self-rated health, and preference of PA. To explore the association between the outcomes, when each health behavior was considered in the dependent variable as an outcome, other health behaviors were included as independent variables.
<sup>b</sup> Generalized linear model with normal distribution and identity link function
<sup>c</sup> Wald test of the interaction term between equivalent household income (ridit score) and survey year

## Discussion

This is the first study to examine the time trend in socioeconomic inequalities in various health behaviors among Japanese adolescents before and during the COVID-19 pandemic. We found a widening of socioeconomic disparities in the achievement of recommended PA and a narrowing of breakfast intake among adolescents. As pertaining to MVPA in particular, while all income groups showed deterioration during the COVID-19 pandemic, it was more prominent in the lowest-income groups. Breakfast intake frequency showed an improving trend among lower-income groups, while it worsened among higher-income groups. For ST, the widening and narrowing was inconclusive. However, it was exacerbated in the higher-income groups. Sleep duration and bowel movement frequency did not change and no socioeconomic disparities were observed. The percentage of the present study participants achieving the recommended sleep duration was poor, at approximately 35%, while other countries achieved roughly 65%.^42^ The prevalence of constipation at about 10% was not relatively high.^7^

Our findings provide deep insights into the widening socioeconomic disparities in PA. The decline in PA among Japanese adolescents during the COVID-19 pandemic was probably a result of the prohibition of extracurricular activities. Japan has a system of extracurricular activities in which teachers are engaged in sports coaching from junior high through high school. The cost of participation in extracurricular activities is mostly small, with minimal expenses for the essential equipment. According to national comprehensive survey data, as of 2019, 66.5% of second-year middle school students were enrolled in sports/exercise programs,^15^ which meant that it played an important role as an equitable opportunity for PA among Japanese adolescents. Local governments adopted measures such as banning extracurricular activities at the municipal level to prevent the transmission of COVID-19. Meanwhile, paid sports organizations provided certain services, such as online lectures. The association between paid sports activities and household income was reported before the pandemic.^43^ It can be presumed that a larger proportion of adolescents from families with sufficient household income were engaged in paid sports and benefited, while those from low-income families not engaged in paid sports were inactive.

Socioeconomic disparities in compliance with breakfast intake recommendation have narrowed: the percentage of lower-income groups has improved, while that of higher-income groups has declined. The reason for the decrease in the percentage of higher income groups is not clear, but it is possible that the overall increase in people’s health awareness due to the pandemic ^44^ led to an improvement in the percentage of low-income groups consuming breakfast. Assuming that the acquisition of knowledge about diet causes behavioral change,^45^ the change to have regular breakfast, irrespective of its content, may be easier than changing other health behaviors. This is because food resources are also abundant in Japan.

A meta-analysis in high-income Western countries found that young people from lower socioeconomic backgrounds have higher levels of ST compared with those from higher socioeconomic backgrounds.^5^ According to our results, in 2019, before the COVID-19 pandemic, the trend in Japan was similar to that in other countries. It can be seen that ST also worsened for higher-income groups due to the pandemic. It is plausible to assume that higher-income groups’ lifestyles were changing as the restrictions on going out led people to spend more time at home. As mentioned above, higher-income groups are more likely to participate in paid private tutoring and sports activities.^43^ Hence, it is assumed that adolescents from higher-income families had more ST at home.

This study had some limitations. First, this was not a longitudinal study. Hence, individual trajectory was unknown. Second, although the sampling method was appropriate to ensure the representativeness of the data, eligible samples were 45.7% and 43.6% of all respondents in 2019 and 2021, respectively. However, the proportion of population in each district/city based on their sizes was similar to that of the nation as a whole. The representativeness of the data would not be deviated considerably. Third, the number of the missing value of income was relatively large. However, the pattern was similar in each survey year. The higher proportion of low income and low PA levels among those excluded from the analysis can be interpreted as an underestimation of the actual association between income and PA in 2019.

This makes it difficult to conclude that the situation has worsened in 2021. However, the necessity of implementing measures to address the socioeconomic disparities in PA is consistent. Although a number of factors related to each health behavior were considered, dietary intake related to bowel movement frequency was only considered for breakfast intake frequency due to lack of data. No socioeconomic disparities in bowel movement frequency were detected previously in Japan ^36^. School lunches may also contribute to reducing the socioeconomic inequality ^10^.

## Conclusion

Our study revealed that economic disparities in the achievement of recommended PA widened, while it narrowed for breakfast intake among adolescents before and during the COVID-19 pandemic. As we found socioeconomic disparities that were insignificant before the pandemic, we recommend the need for urgent supports and medium- and long-term monitoring. Due to the relatively mild restrictions on behavior in Japan compared to other countries (e.g. no lockdowns were imposed),^13^ a more severe situation may occur in other countries. Additional individual-level research is required in various countries andregions as social systems and cultures differ.

## Supporting information

e-Table and e-graph

## Data Availability

All data produced are available online at

https://www.ssf.or.jp/thinktank/sports_life/application/index.html

## Acknowledgements

We would like to thank Editage (www.editage.com) for English language editing.

## Appendices

Appendices for this article are available online.

