## Supplementary material for "Impact of COVID-19 pandemic on the socioeconomic inequality of health behavior among Japanese adolescents: a two-year-repeated cross-sectional survey": e-Table and e-graph

Appendix

e-Table. Missing number and percentage of each variable each survey year.

| | 2019 (N = 1076) | | 2021 (N = 1025) | |
|---|---|---|---|---|
| | n | % | n | % |
| Sex | 0 | 0.0 | 0 | 0.0 |
| Age | 0 | 0.0 | 0 | 0.0 |
| Residence area | 0 | 0.0 | 0 | 0.0 |
| Household income | 239 | 22.2 | 232 | 22.6 |
| Family structure | 0 | 0.0 | 0 | 0.0 |
| Sports participants | 3 | 0.3 | 6 | 0.6 |
| Self-rated health | 4 | 0.4 | 3 | 0.3 |
| Preference of physical activity | 5 | 0.5 | 4 | 0.4 |
| MVPA | 5 | 0.5 | 3 | 0.3 |
| Screen time | 76 | 7.1 | 78 | 7.6 |
| Sleep | 10 | 0.9 | 7 | 0.7 |
| Breakfast | 1 | 0.1 | 3 | 0.3 |
| Bowel movement | 8 | 0.7 | 5 | 0.5 |

*MVPA:* moderate-to-vigorous physical activity

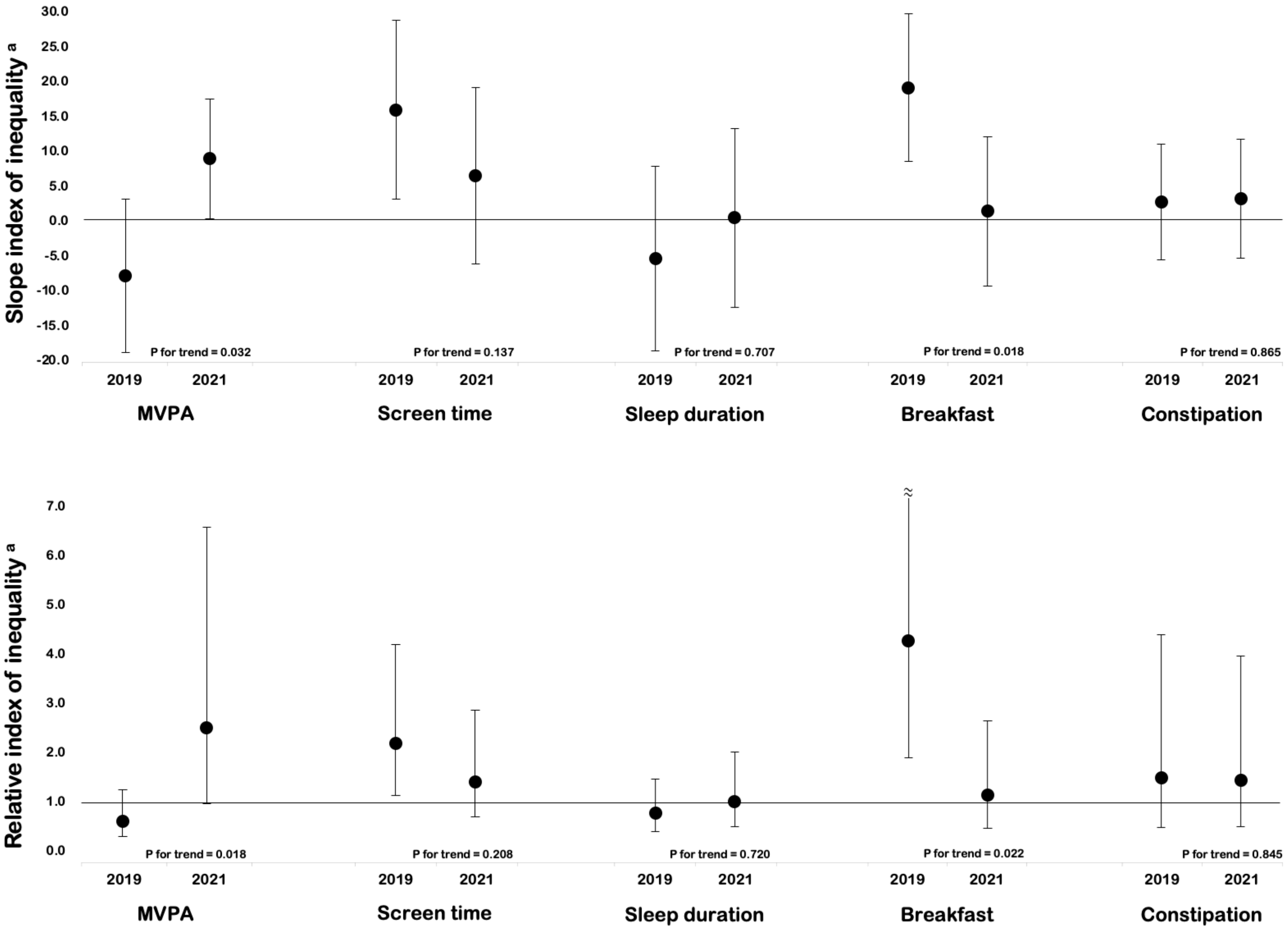


e-Figure. Slope index inequality and relative index of inequality in each health behavior according to the income level indicators by survey year

a Adjusted for place of residence, family structure, sex, age, sports participation, self-rated health, and preference of PA.
